# Tau, β-amyloid, and glucose metabolism following service-related Traumatic Brain Injury in Vietnam war veterans: The AIBL-VETS study

**DOI:** 10.1101/2022.03.10.22272230

**Authors:** Vincent Doré, Tia L. Cummins, Azadeh Feizpour, Natasha Krishnadas, Pierrick Bourgeat, Alby Elias, Fiona Lamb, Robert Williams, Malcolm Hopwood, Victor L. Villemagne, Michael Weiner, Christopher C. Rowe, Alzheimer’s Disease Neuroimaging Initiative, AIBL Research Group

## Abstract

Traumatic Brain Injury (TBI) is common amongst military veterans and has been associated with an increased risk of dementia. It is unclear if this is due to increased risk for Alzheimer’s disease (AD) or other mechanisms. This case control study sought evidence for AD, as defined by the 2018 NIA-AA research framework^1^, by measuring tau, β-amyloid and glucose metabolism using positron emission tomography (PET) in veterans with service-related TBI.

Seventy male Vietnam war veterans — 40 with TBI (aged 68.0±2.5 years) and 30 controls (aged 70.1±5.3 years) — with no prior diagnosis of dementia or mild cognitive impairment underwent β-amyloid (^18^F-Florbetaben), tau (^18^F-Flortaucipir) and ^18^F-FDG PET. The TBI cohort included 15 participants with mild, 16 with moderate, and 9 with severe injury. β-amyloid level was calculated using the Centiloid (CL) method and tau was measured by Standardized Uptake Value Ratios (SUVR) using the cerebellar cortex as reference region. Analyses were adjusted for age and APOE-e4. The findings were validated in an independent cohort from the ADNI-DOD study.

There were no significant nor trending differences in β-amyloid or tau levels or ^18^F-FDG uptake between the TBI and control groups before and after controlling for covariates. The β-amyloid and tau findings were replicated in the ADNI-DOD validation cohort and persisted when the AIBL-VETS and ADNI-DOD cohorts were combined (114 TBI vs 87 controls in total). These findings suggest that TBI is not associated with the later life accumulation of the neuropathological markers of AD.

## Introduction

Military service is associated with an increased risk of traumatic brain injury (TBI), with up to 10% of service personnel experiencing TBI in recent conflicts^2^. TBI has been argued to be a strong environmental risk factor for dementia and epidemiological studies report a 2-4 fold increase in risk amongst veterans with moderate or severe TBI ^3^. TBI has been classified as a modifiable risk factor for dementia^4^. However, the mechanism underlying this increased dementia risk remains unclear. Some studies have reported strong associations between TBI and non-AD forms of dementia but not with AD^5^ or a weaker association between TBI and AD than with vascular dementia and unspecified dementia^6^, while other studies report that TBI is related to higher relative risk of AD ^7-9^ or an earlier age of mild cognitive impairment (MCI) onset^10^. These latter epidemiologic studies have two major limitations; First, they rely on self-report information regarding the history of TBI, and second, the diagnosis of MCI or AD is largely based on medical record review and there is a scarcity of biomarker or direct pathological evidence for this claim.

AD is characterised by dense β-amyloid (Aβ) plaques most abundant in the frontal cortex, the cingulate gyrus, precuneus and lateral parietal and temporal regions ^11^. The second major pathological feature of AD is hyperphosphorylated tau, observed as neurofibrillary tangles (NFTs), found in the transentorhinal areas, limbic and isocortical regions ^12^. Animal studies have demonstrated acute effects of TBI on tau^13-15^, amyloid precursor protein (APP)^16^ and Aβ^17^. It is postulated that these changes in damaged axons contribute to apoptosis and inflammation, indirectly leading to AD^18, 19^. Other mechanistic theories posit that TBI reduces time-to-onset amongst those already at risk of AD, as evidenced by accelerated AD related pathology amongst injured transgenic mice models expressing mutations in APP, and tau^20^. In contrast, enriched levels of phospho-tau species following TBI have been argued to be temporary, returning to baseline levels after just one day ^21, 22^.

Human postmortem studies post TBI are limited and have not provided clear evidence that TBI is associated with Alzheimer’s disease. β-amyloid as diffuse plaques have been found in persons where brain tissue was obtained within hours of severe TBI ^23, 24^. One study of brains with ppostmortemevidence of moderate or severe TBI related to an event ranging from one to 47 years earlier, identified neurofibrillary tangles and β-amyloid plaques in approximately 30% of individuals, with a trend to more extensive distribution than in age matched controls^25^. However, in another postmortem study^26^ on three large community-based cohorts of 1500 brains found no association between TBI with loss of consciousness and AD-like neuropathological features but did find an association with neuropathologic changes related to Lewy body disease and Parkinson’s disease.

Moderate-to-severe TBI survivors, within a year of injury, have been reported to exhibit increased amyloid tracer binding in cortical grey matter and striatum, a pattern of distribution broadly replicating that seen in carriers of mutations in the presenilin-1 (PS1) gene, a driver of early onset AD^27^. In contrast, a study of Aβ deposition with Pittsburgh compound B (PiB) PET in 12 long term survivors of moderate to severe TBI who reported cognitive impairment found no association between tracer uptake and severity of TBI^28^. In the DOD ADNI (www.adni.info.org) dataset, no effect of TBI history on AD biomarkers were found^29,30^.

The current tracers for tau PET detect the 3R/4R form of tau and do not show binding on PET scans in non-AD tauopathies^31^.

Fluorodeoxyglucose (^18^F-FDG) PET allows the measure of hypometabolism, a marker of synaptic/neuronal impairment. Despite inconsistencies in time since injury, TBI severity, age range and injury modality, studies have shown reductions in regional metabolism. A general trend has emerged showing acutely increased glucose utilization in the days after TBI, followed by hypometabolism lasting weeks to months^32-34^. Reductions in regional metabolism have been found in areas of the brain particularly sensitive to TBI, for example in frontal and temporal regions^35-37^, cerebellum, medial temporal lobe, parietal, somatosensory and visual cortices^38, 39^. One group has hypothesised that impaired cerebral glucose metabolism may contribute to promoting amyloidogenesis, abnormal tau hyperphosphorylation and neurofibrillary degeneration, and therefore lead to AD-like pathology^40^. However, the observed depression in glucose metabolism only lasted for a few weeks or months^32^.

The main objective of the Australian Imaging Biomarkers and Lifestyle study of aging - Veterans study (AIBL-VETS) was to investigate with a case-control study design if Vietnam veterans with a history of TBI were more likely to demonstrate the neuropathological markers of AD than controls. Consistent with the 2018 NIA-AA Research Framework for a biological definition of AD^1^, we investigated both β-amyloid and tau biomarkers for AD pathology, and FDG as a marker of neurodegeneration.

## Materials and Methods

The AIBL-VETS study was designed to be compatible with the US Department of Defense funded Alzheimer’s Disease Neuroimaging Initiative veterans study (DOD ADNI) to allow independent validation of findings and pooling of data.

### Participants

Ex-military service personnel, aged 60-85 years, with and without TBI, were recruited through retired veteran organisations such as the Returned Services League and the Vietnam Veterans Association of Australia, as well as the Older Veterans’ Psychiatry Program located at Austin Health, Melbourne, Australia. Exclusion criteria included prior diagnosis of Bipolar Affective Disorder, Schizophrenia, Dementia, Mild Cognitive Impairment, substance abuse/dependence within the last five years, MRI contraindication, major, unstable medical condition, and previous participation in clinical trials involving an amyloid targeting therapy.

To be included in the TBI cohort, participants were required to have sustained at least one TBI between the ages of 16-40 years. TBI severity was assessed based on criteria set by the US Department of Defense and Department of Veterans’ affairs ^41^ (Table 1). Medical records from the time of injury were not available. Given reliance on self-report, and to ensure injuries were given accurate severity ratings (mild/moderate/severe), only participants who were confident on the details of their injury were included. To be included in the control group, participants met the same inclusion/exclusion criteria, but were required to report no prior history of TBI nor Post-Traumatic Stress Disorder (PTSD). Approval for this study was obtained from the Austin Health Human Research Ethics Committee, the US Human Subjects Research Protection Office of the US Army Medical Research and Material Command, and the Australian Department of Veterans Affairs Ethics Committee. All participants provided consent prior to participating, and there were no direct incentives offered for participation.

**Table 1:** Criteria used by Department of Defense and Veterans Affairs (VA/DoD) to categorize head injury severity

|  | Mild | Moderate | Severe |
| --- | --- | --- | --- |
| Loss of consciousness | 0 - 0.5 hrs | 0.5 - 24 hrs | >24 hrs |
| Alteration of consciousness | A moment - 24 hrs | >24 hrs; severity based on other criteria | >24 hrs; severity based on other criteria |
| Post-traumatic amnesia | 0 - 1 day | 1 - 7 days | >7 days |
| Glasgow Coma Scale | 13 - 15 | 9 - 12 | 3 - 8 |
| Structural Imaging | Normal | Normal/abnormal | Normal/abnormal |

### Procedure & Materials

All participants were screened over the phone to ensure they matched study criteria. Those deemed suitable for the initial assessments were invited into the research center to undergo psychiatric and neuropsychological assessment and an interview to obtain detailed TBI history. Participants were asked to give a detailed account of events surrounding the injury, including age at injury, injury cause, presence and length of unconsciousness, alteration of consciousness and post traumatic amnesia, as well as information as to medical attention sought, and disruption of usual activities due to injury. Based on this information, and in relation to the information included in Table 1, each injury was classified as mild, moderate, or severe.

#### Psychiatric wellbeing

The psychiatric evaluation consisted of several measures to assess PTSD severity, drug and alcohol use, sleep quality and medical history. A PTSD diagnosis was allocated based on the Clinician Administered PTSD Scale (CAPS)^42^ lifetime and current score. A lifetime CAPS score of over 40 was indicative of previous PTSD, whilst a current CAPS score of over 40 indicated current PTSD. The Addiction Severity Index-lite^43^ was used to assess alcohol/substance use, and the Pittsburgh Sleep Quality Index (PSQI) ^44^, sleep quality and disturbance. A score >5 on the PSQI was indicative of poor sleep quality.

#### Other measures

Participants also self-reported years of education, military-service history, cigarette smoking status and medical history. Comorbidities such as hypertension, ischemic heart disease, stroke, diabetes, migraine, and sleep apnoea were determined via self-report in an interview with the study psychiatrist. Apolipoprotein E (APOE) genotype was determined by direct sequencing. The Wechsler Test of Adult Reading (WTAR) was also assessed to provide a measure of premorbid intelligence. Participants also completed the Combat Exposure Scale (CES)^45^ to classify the level of wartime stressors experienced.

#### Image acquisition and processing

Participants underwent tau, amyloid and ^18^F-FDG PET, performed on separate days using ^18^F-Flortaucipir, ^18^F-Florbetaben, and ^18^F-FDG respectively. All radiotracers were produced on site, at Austin Health. The scans were acquired on a Siemens 128 mCT PET/CT camera at the University of Melbourne. A 30-minute acquisition was performed 75-minutes post-injection of ^18^F-Flortaucipir, and 20 minutes scans were acquired 90 minutes post injection of ^18^F-Florbetaben and after 30 minutes uptake time while resting quietly in a dimly lit room post-injection of ^18^F-FDG. Low dose CT scan was used for attenuation correction. There was no correction for partial volume effects. In a previous paper, we reported no difference in hippocampal or grey matter volumes between the TBI and control groups in this cohort^46^.

##### DOD ADNI validation cohort

Data and scans from the US Department of Defense Alzheimer’s Disease Neuroimaging Initiative (DOD ADNI) database (adni.loni.usc.edu) were used to validate the Aβ and tau PET findings. The primary goal of ADNI has been to test whether serial magnetic resonance imaging (MRI), PET, as well as other biological, clinical, and neuropsychological assessment can be combined to detect and measure the progression of preclinical, mild cognitive impairment (MCI) and early Alzheimer’s disease. For up-to-date information see www.adni.info.org. In the DOD ADNI cohort, 57 normal control (NC) and 74 TBI (16 mild TBI, 58 moderate-to-severe TBI) cognitively unimpaired veterans underwent a ^18^F-Florbetapir β-amyloid scan while 30 NC and 46 TBI (11 mild, 35 severe) had a ^18^F-Flortaucipir tau scan (Supplementary Table 1). After download, these scans were processed using the methods described below for the Australian veteran cohort to provide compatible Centiloid and SUVR results.

#### Image analysis

Reconstructed PET images were processed using CapAIBL, a previously validated tracer uptake quantification software package^47^. The standardised uptake value ratio (SUVR) quantification process is described in-depth elsewhere ^48^, however, in brief, an adaptive atlas was automatically fitted to each Aβ PET image to match the Aβ PET retention pattern. Each image was then spatially normalised to the best fitting atlas. Centiloid values were then computed using the SPM8 mask and CapAIBL calibration equations from Bourgeat, Doré et al. (2018)^49^.

All tau and ^18^F-FDG scans were normalised using the same CapAIBL software, but instead of the Aβ analysis, multi-tau atlases^47^ and a mean ^18^F-FDG atlas were used. Spatially normalised tau scans were sampled in three different composite regions, as per earlier work^50^: Mesial-temporal (Me), Temporoparietal (Te), and rest of neocortex (R). The Me region comprised entorhinal cortex, hippocampus, parahippocampus and amygdala, and the Te region comprised regions inferior and middle temporal, fusiform, supramarginal and angular gyri, posterior cingulate/precuneus, superior and inferior parietal, and lateral occipital. Rest of neocortex (R) was made up of dorsolateral and ventrolateral prefrontal, orbitofrontal, gyrus rectus, superior temporal and anterior cingulate. Glucose metabolism was also investigated in the neocortex region (a composite region of frontal (dorsolateral, ventrolateral, and orbitofrontal), parietal (superior parietal and precuneus), lateral temporal (superior, middle, and inferior), lateral occipital lobe, gyrus supra-marginalis, gyrus angularis and anterior and posterior cingulate.), in the frontal, the Me and in the posterior cortical index (PCI) comprising the lateral temporal, parietal, and precuneus cortex.

For FTP and FDG, Standardised Uptake Values (SUV) were calculated for all brain regions examined. The Standardised Uptake Value Ratio (SUVR) was the primary outcome variable for the PET assessments and was generated by dividing the regional SUVs by the cerebellar cortex SUV.

#### Visual reading of images

All ^18^F-Florbetaben, ^18^F-Flortaucipir and FDG images were also visually inspected. Each image was assessed by a nuclear medicine specialist experienced in interpretation of these scans. All ^18^F-Florbetaben and FDG scans were rated as *positive* or *negative*, and ^18^F-Flortaucipir images were rated as either *negative, equivocal*, or *positive*.

### Data analysis

Power analysis was calculated from amyloid scans in age matched healthy controls from the Australian Imaging Biomarkers Lifestyle study of aging (AIBL) and showed that to observe a moderate effect size between groups (Cohen’s d ≥ 0.5), with 80% power, a total of 62 participants would be required. Percentages were calculated for categorical variables. Participant characteristics and demographics were compared between individuals in the TBI cohort and the veteran control group using t -tests and the Wilcoxon signed rank test (when variables were not normally distributed) for continuous variables and Fisher’s exact test for categorical variables, as well as Cohen’s d to measure effect sizes. Hierarchical regressions were used to investigate the influence of covariates, including age, APOE e4 status, years of education, premorbid intellectual functioning, and lifetime PTSD severity. The Weschler Test of Adult Reading was used to provide an estimate of IQ, and CAPS Lifetime score as a measure of lifetime severity of PTSD. Variables that had a significant effect on the outcome variable were then controlled for in an analysis of covariance (ANCOVA). A p-value of less than 0.05 was deemed statistically significant.

## Results

### Demographics

When compared to the control group, participants with a TBI had significantly fewer years of education, lower levels of premorbid intellectual functioning, higher Body Mass Index (BMI), higher PTSD, depression and distress scores, were more likely to endorse previous diagnosis of sleep apnoea and more likely to carry the APOE-e4 allele (Table 2). Of the 40 TBI subjects, 12 met criteria for current PTSD. Of the TBI cohort, 15 participants had suffered mTBI, 16 moderate, and 9 severe. The 3 TBI groups did not differ from each other in terms of demographics or medical comorbidities. Injuries were sustained from a variety of mechanisms and further details are included in Figure 1. The most common cause of injury, across all severities, was sports related, followed by motor vehicle accidents. The average age at injury was 24.2 (±5.5), with the average time since injury 44.1 (±5.3) years. The range for time since injury was 30-53 years.

**Table 2:**
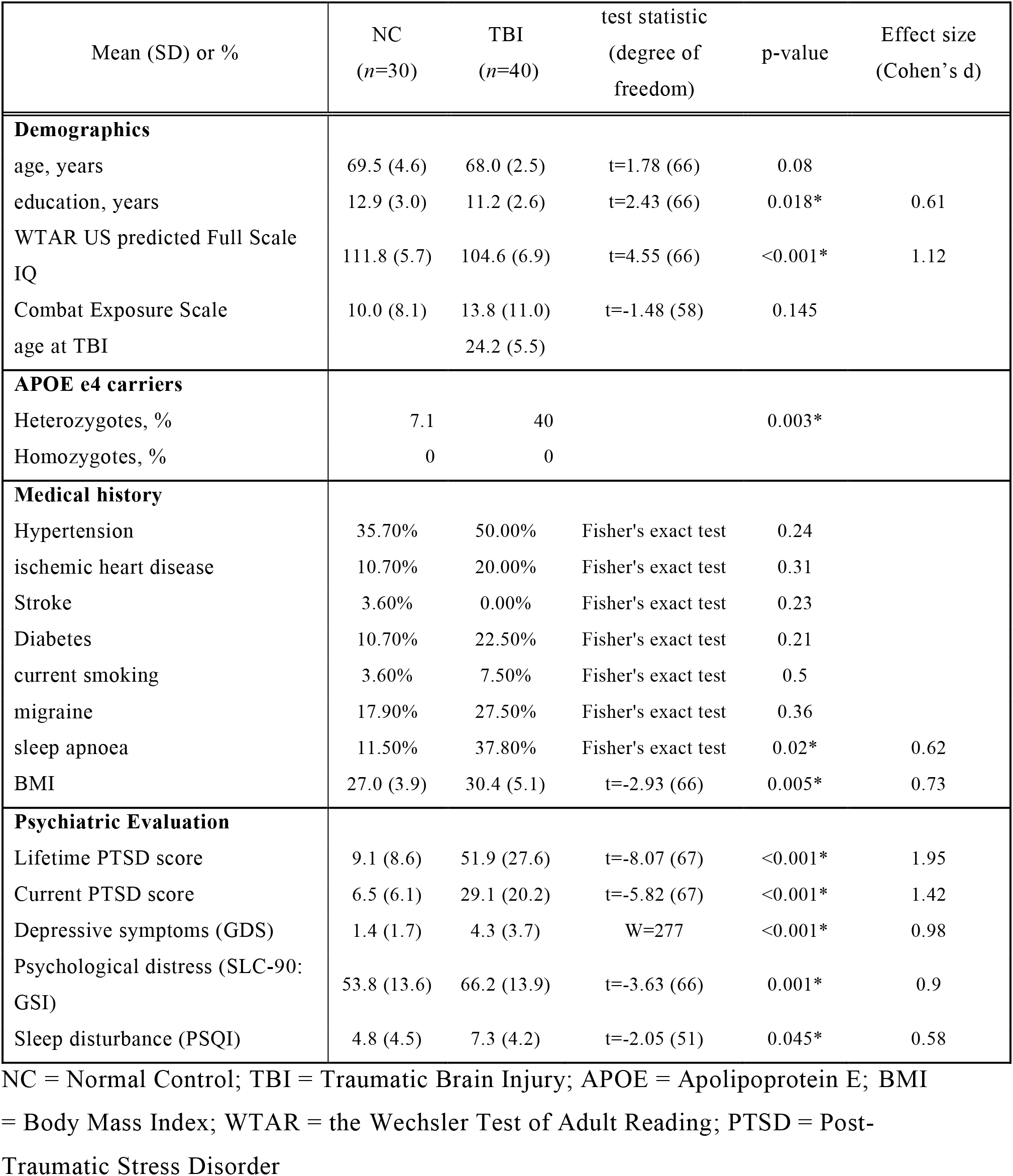
AIBL-VETS Participant demographics

**Figure 1:**
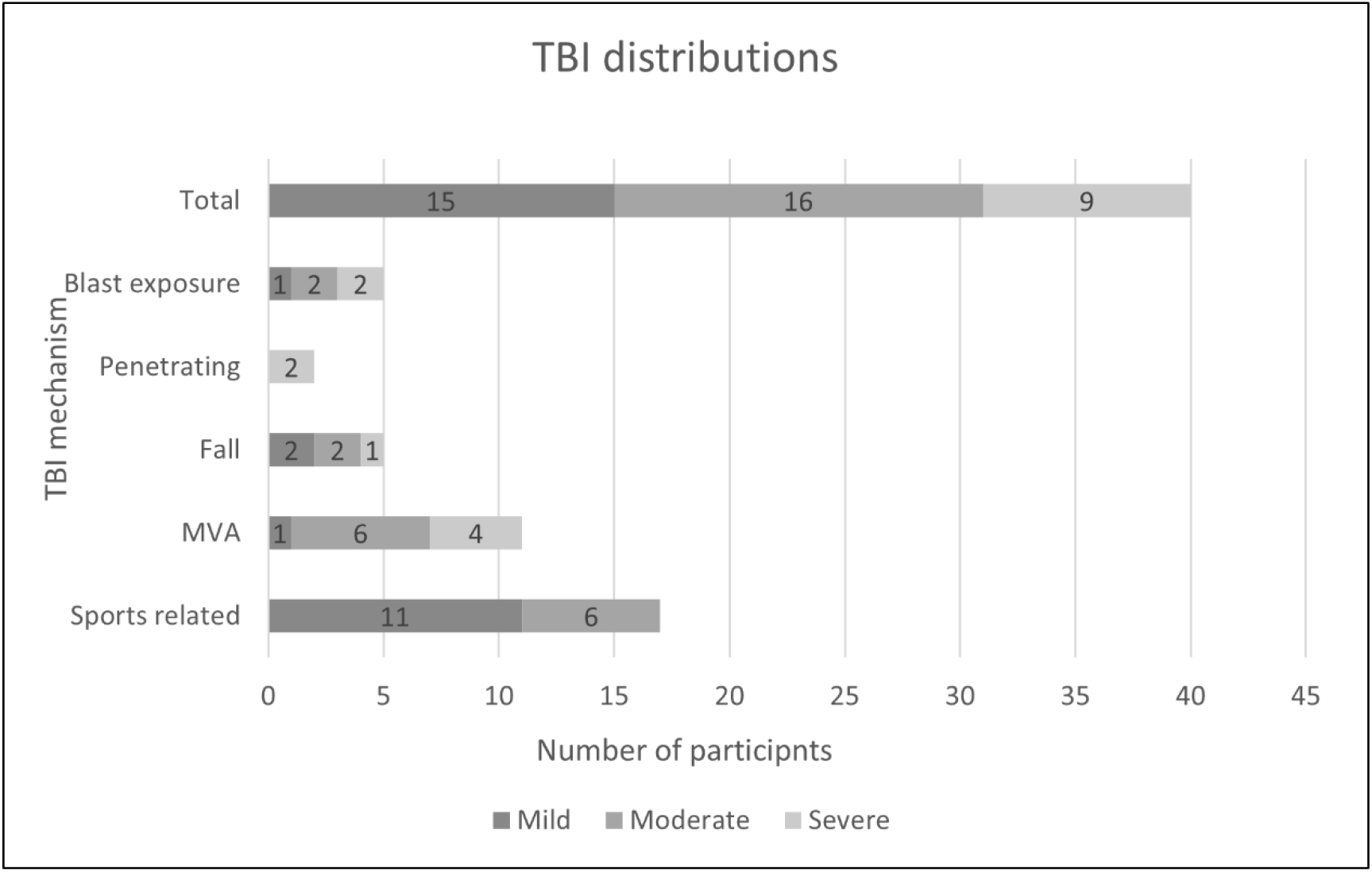
TBI characteristics. MVA – motor vehicle accident

### Positron Emission Tomography

After examining demographic, medical, and psychiatric covariates, results from a series of hierarchical linear regressions showed that only APOE had significant association with ^18^F-Florbetaben SUVR. We have previously reported that PTSD was not associated with PET findings in this AIBL-VETS cohort^58^. A T-test on ^18^F-Florbetaben SUVR showed no difference between TBI and NC (Cohen’s d=0.31, p=0.2), despite the larger number of APOE4 carriers in the TBI group. When considering only the APOE non-E4 carriers, the Cohen’s d was 0.21 (p=0.44). After controlling for APOE using an ANCOVA, the difference was even smaller between TBI and the NC cohort in ^18^F-Florbetaben SUVR (F=0.33, p>0.55). Similar results were found in the DOD ADNI cohort and when merging the AIBL-VETS and DOD ADNI cohorts (Figure 2 B and C).

**Figure 2:**
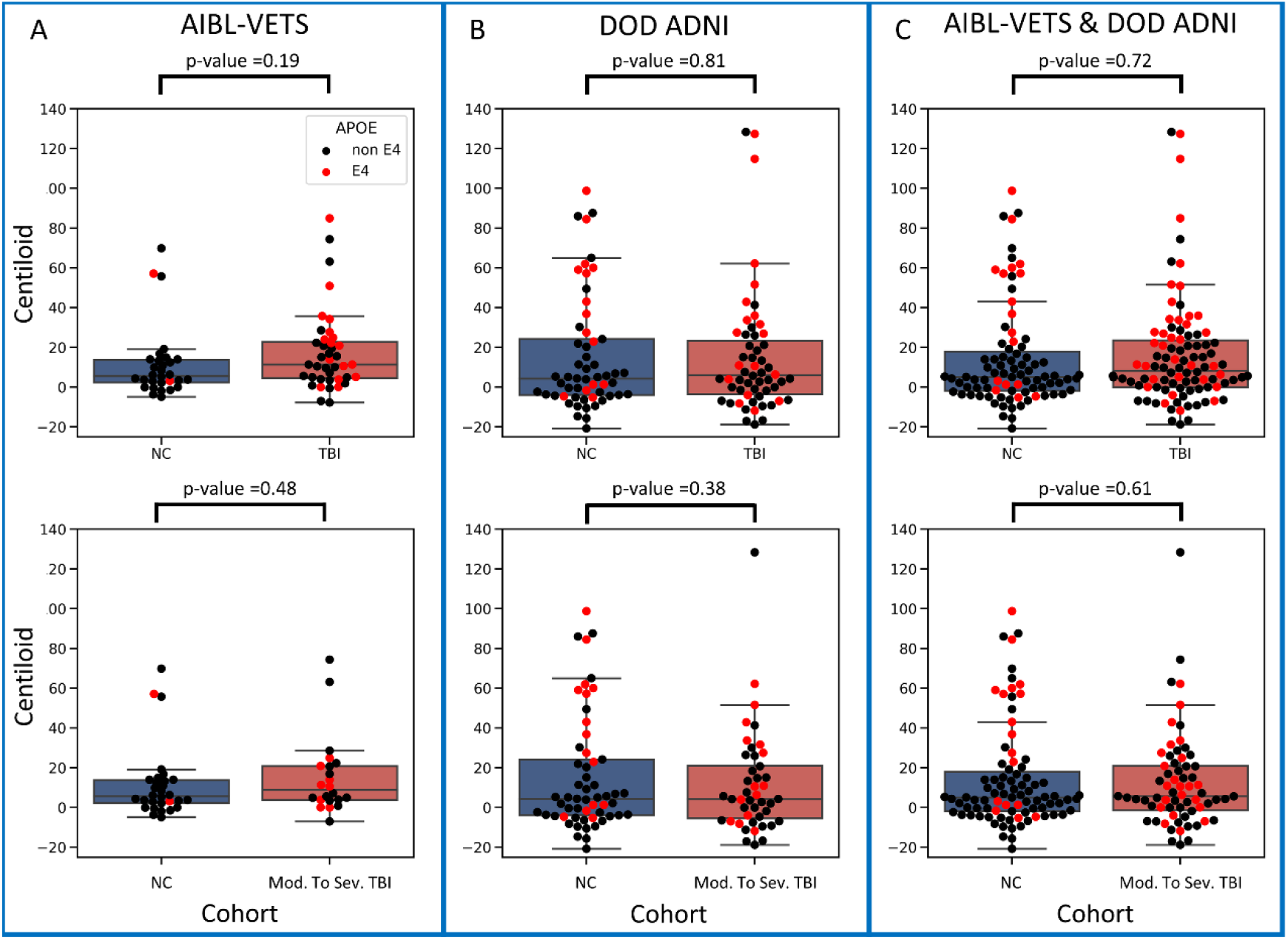
^18^F-Florbetaben SUVRs for NC and TBI groups from AIBL-VETS and DOD ADNI cohorts and both cohorts together. Top row – normal controls (NC) vs all TBI; Bottom row– NC vs moderate or severe TBI

T-tests and ANCOVA on global and regional ^18^F-Flortaucipir showed no significant difference between controls and TBI subjects from the AIBL-VETS cohort (Supplementary Figure 1), from the DOD ADNI cohort (Supplementary Figure 2) and from the merging of both cohorts (Figure 3).

**Figure 3:**
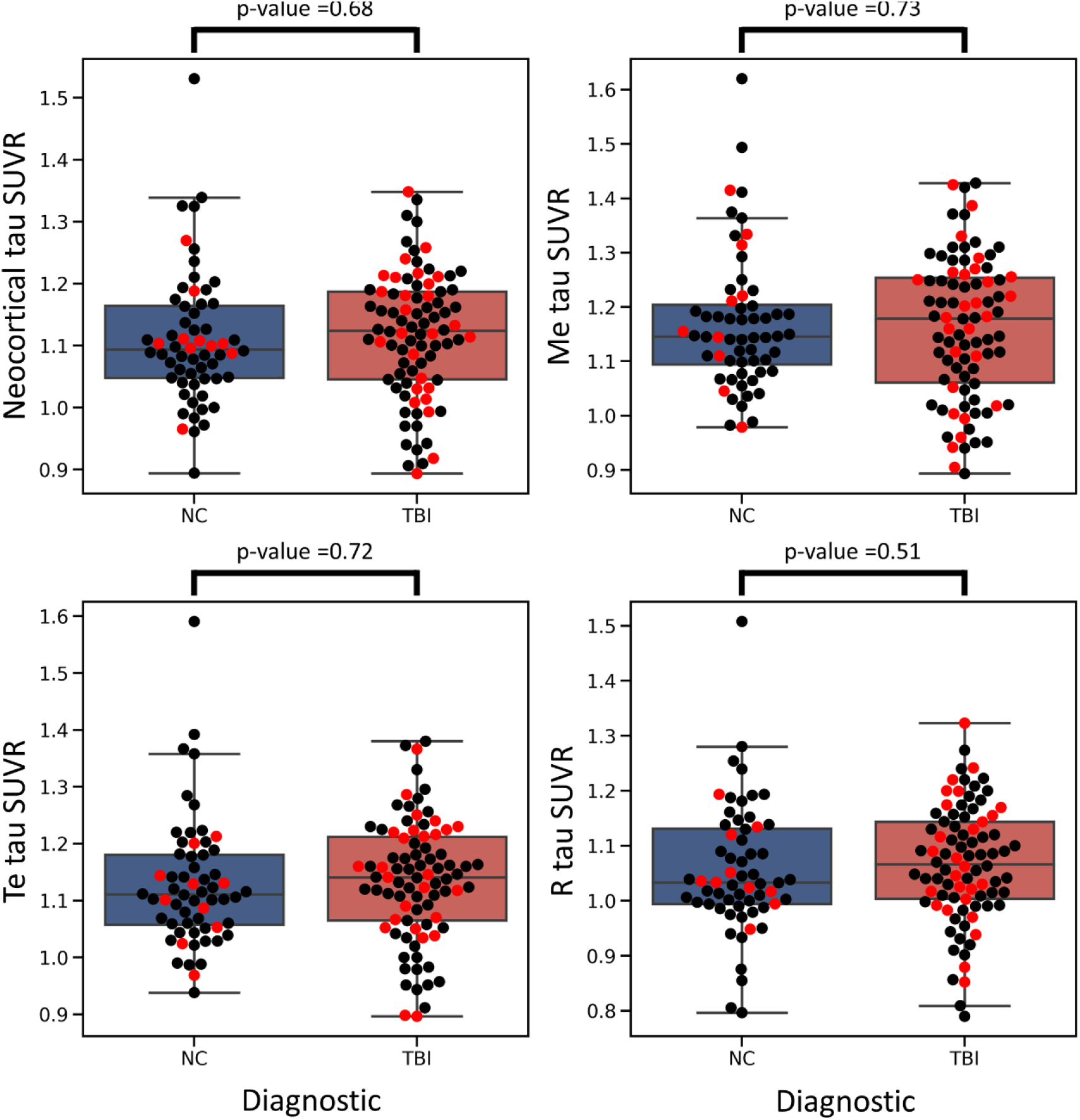
^18^F-Flortaucipir SUVRs for NC and TBI groups from AIBL-VETS and DOD ADNI cohorts merged together. Me – Mesial-temporal; Te – Temporal parietal; R - Rest of neocortex.

We found no significant difference in the ^18^F-FDG SUVs from the cerebellum cortex between the two groups, as reported in previous studies^51^. This allowed us to analyse the ^18^F-FDG SUVRs. No significant differences were found in the global and regional quantification of FDG uptake between NC and TBI groups (Figure 4).

**Figure 4:**
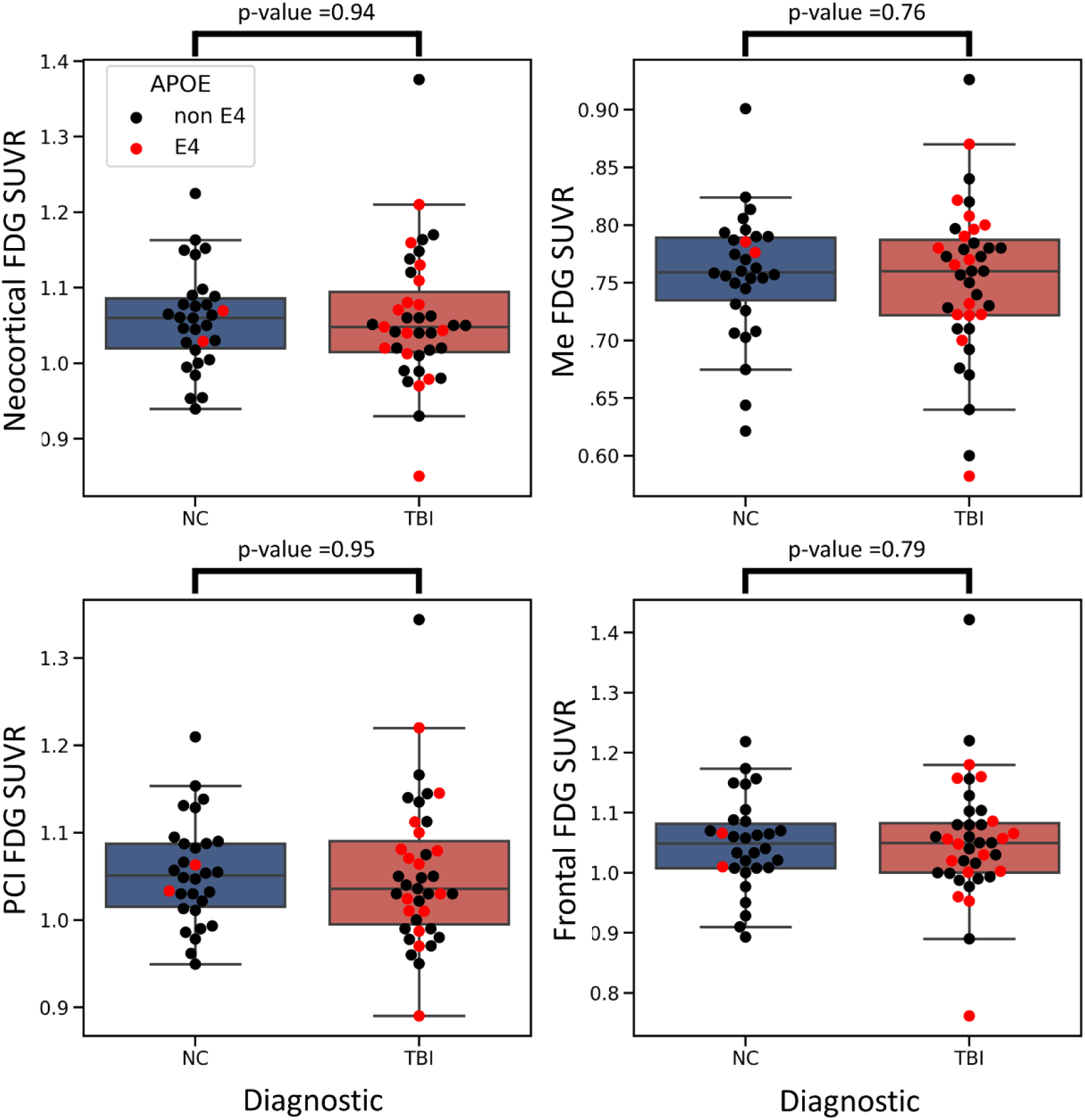
^18^F-FDG SUVRs for NC and TBI groups in AIBL-VETS. Me – Mesial-temporal; PCI – Posterior Cortical Index; Te – Temporal parietal; R-Rest of neocortex.

### Effect of injury severity

To investigate the impact of injury severity, the TBI cohort was subdivided into mTBI (n=15) and moderate-to-severe TBI (n=25) and we performed the analysis between NC and the moderate to-severe TBI (Figure 1, lower row). We did not find any significant covariate and there were no significant differences between NC and moderate-to-severe TBI in centiloid values nor in tau SUVR (Supplementary Figure 3). Cohen’s d was of the same order as the previous analysis (Cohen’s d>-0.30).

When visual classifications were analysed, no differences in Aβ or tau positivity rates were found between TBI and control cohorts, even after removing the mild injuries from the analysis.

## Discussion

In this study, we employed amyloid, tau and ^18^F-FDG PET, and APOE genotyping to investigate the presence of the neuropathological hallmarks of AD in a homogenous cohort of veterans three-to-five decades after TBI. The current study did not find evidence of increased amyloid deposition amongst veterans with TBI, even in those with more severe injuries. Our findings were replicated in the DOD ADNI validation cohort. This result is in line with previous studies investigating the association between TBI and AD pathology with post-mortem data^26, 52^ and with amyloid PET imaging^28, 30^. Furthermore, recent findings in clinically normal older adults confirm that in a longitudinal Aβ-PET study (two Aβ-PET scans, 0.5 to 4 years apart), adults with mTBI did not have a significantly higher rate of Aβ accumulation over time than those with no remote head trauma^53^. Several other studies have reported association between TBI and amyloid deposition^24, 27^ however time between TBI and in-vivo amyloid assessment was less than 1 year, or the association did not reach significance^25^.

Using the MeTeR scale, the current study investigated tau accumulation in AD vulnerable regions, nevertheless no significant differences in tracer uptake were found on two different veteran cohorts (AIBL-VETS and DOD ADNI), even amongst those with moderate-to-severe injuries. In contrast to our findings, a study using ^11^C-PBB3 tau tracer found that patients with mild-repetitive or severe TBI had higher ^11^C-PBB3 binding in the neocortical grey and white matter than healthy control participants ^54^. Also in one post-mortem study, a higher prevalence of subjects with NFT among TBI was reported but this was significant only in TBI subjects younger than 60 years old^25^. No difference was observed in a subsequent, much larger cohort^26^.

We also report no difference in ^18^F-FDG uptake. Previous studies reported low glucose metabolism between several days and a few months after TBI^55^. However, it was also reported that the depression in glucose metabolism subsided after a few months ^32^.

In the 2020 Lancet report on dementia prevention^4^, TBI was listed as a factor that can contribute to increased dementia risk. However, no differentiation between AD and non-AD dementia was performed. The normal level of Aβ deposition, tau aggregation and glucose metabolism in TBI subjects compared with normal matched individuals does not support the premise that TBI can be a risk factor for Alzheimer’s Disease. This suggests that the observed increase in dementia risk may be due to other causes of dementia as has been suggested, such as Dementia with Lewy Bodies^26^ or frontotemporal dementia (FTD)^56, 57^.

Low brain resilience or cognitive reserve leading to earlier clinical manifestation of dementia for a given degree of neuropathology could also contribute to the consistently reported increase in the prevalence of dementia in those with history of TBI. The prevalence of AD is partially censored by deaths from other causes in the elderly so earlier onset in those with TBI would also suggest higher prevalence. The AIBL-VETS TBI cohort had many factors that could create low brain resilience or reserve. These include diminished white matter integrity in those with moderate or severe TBI as previously reported by the authors^46^. The TBI cohort also scored significantly lower on the WTAR measure of premorbid intellectual functioning than controls and as expected, higher on a number of psychiatric measures. These included measures of anxiety, somatization, obsessive compulsive behaviours, interpersonal sensitivity, depression, hostility, paranoid ideation, psychoticism, in addition to lifetime and current PTSD symptomology. Other health measures indicated that the TBI cohort have a significantly higher BMI than controls, which may account also for the higher self-reported incidence of sleep apnoea in this group. Prevalence of ischemic heart disease and diabetes, whilst not significantly different to controls, was nearly doubled in the TBI cohort.

The results from the current study provide a unique perspective into the long-term effects of TBI. Strengths of the study include: the reasonably large imaged cohort (114 TBI and 87 veteran controls when AIBL and ADNI cohorts were merged), homogenous TBI and control participants by restricting the study to veterans of the Vietnam war, only including veterans three-to-five decades after injury, and using a range of biomarkers to investigate the presence of neuropathological markers of AD. Our analysis was validated with the independent DOD ADNI cohort.

Our study has some limitations. Medical records were not available to confirm TBI severity, therefore, the study team were reliant upon self-report, which may have led to an under or over-estimation of injury severity. In addition, sample sizes, while large for a TBI PET study, restricted group separation by injury mechanism. This resulted in a mixture of single and repetitive injuries in the mTBI group, and blast, penetrating and other injury mechanisms amongst the moderate-to-severe TBIs. It was not possible to exclude participants with TBI in addition to PTSD, and this may limit the applicability of these findings to a number of other TBI cohorts.

In summary, the findings of this study suggest that TBI is not associated with the later life accumulation of the neuropathological markers of AD.

## Data Availability

Anonymized data from AIBL-VETS cohort may be shared upon request to the corresponding author from a qualified investigator, subject to restrictions according to participant consent and data protection legislation.

http://adni.loni.usc.edu/data-samples/access-data/

## Acknowledgements

Some of the data used in the preparation of this article was obtained from the Australian Imaging Biomarkers and Lifestyle flagship study of aging (AIBL), and the Australian Dementia Network (ADNeT) that receive funding support from the National Health and Medical Research Council (NHMRC).

Data collection and sharing for this project was also funded by the Alzheimer’s Disease Neuroimaging Initiative (ADNI) (National Institutes of Health Grant U01 AG024904) and DOD ADNI (Department of Defense award number W81XWH-12-2-0012). ADNI is funded by the National Institute on Aging, the National Institute of Biomedical Imaging and Bioengineering, and through generous contributions from the following: AbbVie, Alzheimer’s Association; Alzheimer’s Drug Discovery Foundation; Araclon Biotech; BioClinica, Inc.; Biogen; Bristol-Myers Squibb Company; CereSpir, Inc.; Cogstate; Eisai Inc.; Elan Pharmaceuticals, Inc.; Eli Lilly and Company; EuroImmun; F. Hoffmann-La Roche Ltd and its affiliated company Genentech, Inc.; Fujirebio; GE Healthcare; IXICO Ltd.;Janssen Alzheimer Immunotherapy Research & Development, LLC.; Johnson & Johnson Pharmaceutical Research & Development LLC.; Lumosity; Lundbeck; Merck & Co., Inc.;Meso Scale Diagnostics, LLC.; NeuroRx Research; Neurotrack Technologies; Novartis Pharmaceuticals Corporation; Pfizer Inc.; Piramal Imaging; Servier; Takeda Pharmaceutical Company; and Transition Therapeutics. The Canadian Institutes of Health Research is providing funds to support ADNI clinical sites in Canada. Private sector contributions are facilitated by the Foundation for the National Institutes of Health (www.fnih.org). The grantee organization is the Northern California Institute for Research and Education, and the study is coordinated by the Alzheimer’s Therapeutic Research Institute at the University of Southern California. ADNI data are disseminated by the Laboratory for Neuro Imaging at the University of Southern California.

We also thank the participants who took part in the study and their families.

## Authorship Contribution Statement

**Vincent Doré:** Methodology, Formal analysis, Writing - Original Draft, **Tia L. Cummins:** Data Curation, Methodology, Formal analysis, Writing - Original Draft, **Azadeh Feizpour:** Writing - Original Draft, **Natasha Krishnadas:** Writing - Review & Editing, **Pierrick Bourgeat:** Formal analysis, **Alby Elias:** Data Curation, **Fiona Lamb:** Data Curation, **Robert Williams:** Data Curation, **Malcolm Hopwood:** Writing - Review & Editing, **Victor V. Villemagne:** Writing - Review & Editing, **Michael Weiner:** Writing - Review & Editing, **Christopher C. Rowe:** Conceptualization, Resources, Writing - Review & Editing

