## Supplementary Table 1 for "Tau, β-amyloid, and glucose metabolism following service-related Traumatic Brain Injury in Vietnam war veterans: The AIBL-VETS study"

**Supplementary Table 1**: ADNI-DOD Participant Demographics

|  | **NC** | **TBI** | **Effect size (Cohen’s d)** | **P-value** |
| --- | --- | --- | --- | --- |
| **Sample #** | 57 | 74 | NA | NA |
| **Age** | 71.5 (6.0) | 68.8 (9.1) | 0.34 | 0.06 |
| **APOE (85% completed)** | 26.7% | 32.1% | χ=0.17 | 0.67 |
| **Education, years** | 16.0 (2.2) | 15.3 (2.3) | 0.29 | 0.1 |
| **TBI (age)** | NA | 24.1 (12.2) | NA | NA |
| **MMSE** | 28.9 (1.0) | 28.3 (1.6) | 0.41 | 0.02* |
| **CDR** | 0.02 (0.25) | 0.03 (0.3) | -0.03 | 0.8 |

APOE = Apolipoprotein E; TBI = Traumatic Brain Injury; MMSE = Mini-Mental State Examination; CDR = Clinical Dementia Rating
