## Supplementary Figure 2 for "Tau, β-amyloid, and glucose metabolism following service-related Traumatic Brain Injury in Vietnam war veterans: The AIBL-VETS study"

**Supplementary Figure 2:** ^18^F-Flortaucipir SUVRs for NC and TBI groups from the ADNI-DOD cohort. Me – Mesial-temporal; Te – Temporal parietal; R - Rest of neocortex.

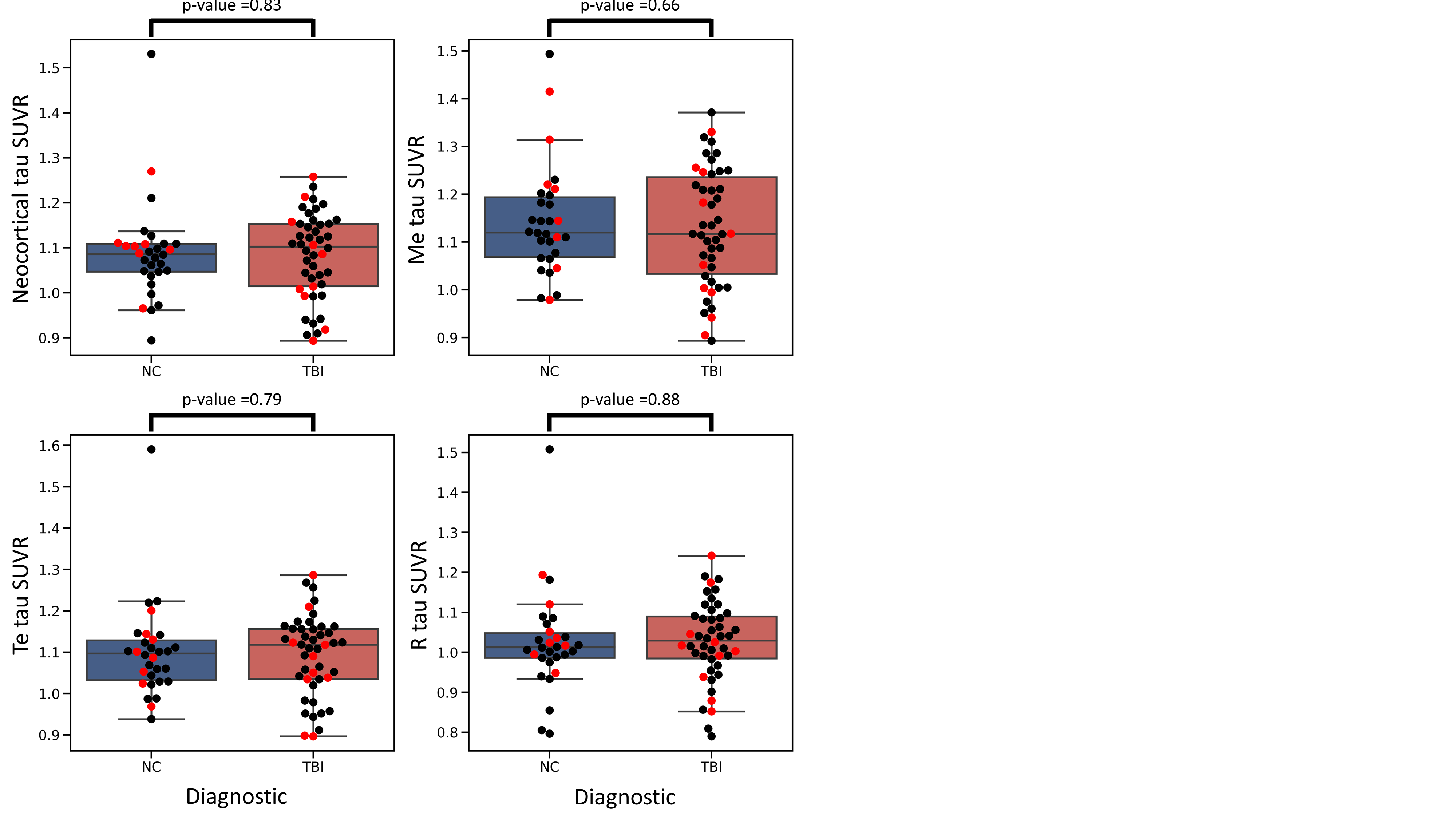
